# Fit-for-purpose bioanalytical validation of a three-cytokine human PBMC ELISpot panel (IFN-γ, IL-2, IL-4) using CEFX and CPI antigen pools for clinical trial immunomonitoring under a GLP-aligned quality system

**DOI:** 10.64898/2026.11.22.26368554

**Authors:** Kaitong Li, Min Tang, Xiaoliang Zhai, Hongxu Cao, Peng Guo, Ning Zhang, Pengyang Fan, Lijie Jiang, Zhenyuan Cui, Hongjuan Wang, Zhuo Wang

## Abstract

Cellular immunity is an increasingly required endpoint in clinical trials of vaccines, immuno-oncology agents, and cell therapies. Multi-cytokine validation packages under a single quality system remain uncommon, and the permissible interval between blood draw and plating or freezing is rarely demonstrated experimentally. We performed GLP-aligned, fit-for-purpose validation of three human peripheral blood mononuclear cell (PBMC) ELISpot assays, following Global CRO Council recommendations and FDA, NMPA, and CFDI guidance. IFN-γ and IL-2 were stimulated with CEFX Ultra SuperStim Pool (24 h each, plus a validated 48 h IFN-γ regimen) and IL-4 with CPI Antigen Pool (CTL-CPI-001; 48 h) throughout all validation arms. We characterized limit of detection (LOD), lower limit of quantification (LLOQ), precision, linearity across six plating densities, robustness, pre-analytical PBMC functional stability under five delayed-processing conditions in fresh and cryopreserved arms. All assays met predefined criteria (≥80% of samples with %CV ≤40% above LLOQ; R^2^ ≥0.97); LOD/LLOQ were 13/20 and 37/45 SFU/well for IFN-γ at 24 h and 48 h, 9/27 for IL-2, and 4/27 for IL-4. Freshly plated PBMC remained within acceptance after up to 8 h plus 24 h of ambient holding, whereas cryopreservation after prolonged delay failed at 8+16 h for IL-2 and IL-4 and at 8+24 h for all three assays, driven by post-thaw viability <80%. These data define evidence-based sample-handling windows for multicenter trials. The resulting three-cytokine platform is fit-for-purpose for regulated clinical immunomonitoring.

## Introduction

Cellular immunity is now a pivotal endpoint in the development of vaccines, anti-infective interventions, immuno-oncology agents, and cell and gene therapies. Unlike humoral readouts, functional T-cell assays capture the frequency and quality of antigen-responsive cells, and they are increasingly required by regulators and sponsors as complementary or primary pharmacodynamic and immunogenicity measures (1-3). Among the available platforms, the enzyme-linked immunospot (ELISpot) assay retains a distinctive position: it enumerates individual cytokine-secreting cells at single-cell resolution, requires relatively small numbers of cells, tolerates batched cryopreserved samples, and scales to the sample volumes of multicenter trials. Compared with intracellular cytokine staining, ELISpot offers higher throughput and simpler instrumentation; compared with soluble cytokine ELISA or multiplex bead assays, it reports responder-cell frequency rather than bulk secreted protein, which is often the biologically interpretable quantity in immunomonitoring (1-3). Standardization of plate reading and counting, including automated evaluation and harmonized counting gates, has further improved reproducibility across laboratories (2,4).

IFN-γ ELISpot is the dominant readout for type 1 cellular responses and has been validated for viral antigens, CMV, influenza, and SARS-CoV-2 spike in both human and non-human primate settings (4,5-7). IFN-γ alone, however, provides an incomplete picture of T-cell biology. IL-2 is produced earlier after activation and is associated with proliferative capacity, memory differentiation, and T-cell help; its co-measurement with IFN-γ distinguishes polyfunctional from narrowly differentiated responses. IL-4 provides a Th2-axis counterbalance that is informative when balanced immune profiling is required, for example when evaluating Th1/Th2 skewing, hypersensitivity risk, adjuvant effects, or vector- and product-specific immune deviations. Formal validations of dual-color IFN-γ/IL-4 ELISpot and IFN-γ/IL-2 FluoroSpot assays have demonstrated that multi-cytokine spot formats can satisfy bioanalytical acceptance criteria for clinical use (8,9), and optimized cultured ELISpot/FluoroSpot protocols continue to expand the methodological toolkit (10). What remains comparatively scarce are GLP-aligned, fit-for-purpose validation packages that cover three single-color cytokine assays under one laboratory quality system and that report extensive pre-analytical PBMC handling data alongside analytical performance.

For regulated bioanalysis, ELISpot validation is guided by industry consensus rather than a single dedicated regulatory guideline. White papers from the Global CRO Council (GCC) and the WRIB collaboration summarize recommended parameters — LOD, LLOQ, precision, specificity and controls, linearity/range, robustness, stability, and critical-reagent management — and frame acceptance around coefficients of variation of ≤30–40% for cell-based assays, with population-level rules such as ≥80% of samples meeting the precision criterion (11,12). The FDA immunogenicity guidance provides analogous fit-for-purpose principles for assay development and validation (13), while regional requirements such as the Chinese Good Laboratory Practice regulations and the 2025 guideline for bioanalytical laboratories in drug clinical trials govern study conduct and data integrity at the testing facility (14,15). Translating these recommendations into an operational clinical assay also demands critical-reagent management (including new-lot bridging) and realistic sample logistics, because multicenter trials rarely achieve immediate PBMC isolation.

Pre-analytical handling is arguably the most under-reported source of variability in cellular immunomonitoring. Blood is commonly collected at sites without cell-processing capability, shipped at controlled ambient temperature, and processed after delays of several hours to more than a day; cryopreservation is then used to batch samples for a single analytical run. Delayed processing can alter granulocyte contamination, cell viability, and antigen-specific responsiveness, and a method optimized on freshly isolated PBMC is not automatically valid for delayed or cryopreserved material (3,5,10). Yet most published validation reports characterize analytical performance only, leaving the permissible window between blood draw and plating or freezing to be assumed rather than demonstrated. GCC recommendations explicitly call for SOP content on sample collection and handling times and on critical reagents (11); empirical data defining those windows are therefore of direct practical value.

Here we report formal bioanalytical validation of three human PBMC ELISpot assays performed under GLP at a single bioanalytical laboratory: IFN-γ and IL-2 stimulated with CEFX Ultra SuperStim Pool, and IL-4 stimulated with CPI Antigen Pool (CTL-CPI-001). We validated LOD, LLOQ, precision, linearity, robustness, and PBMC functional stability across five defined handling conditions, and we additionally report partial validation supporting CEFX new-lot bridging for IFN-γ. Our objective was to establish a deployable, quality-system-controlled three-cytokine platform with analytically and logistically defined operating boundaries for clinical cellular immunity monitoring.

## Materials and Methods

### Study design and compliance

Method validation studies were conducted at JOINN Medical Testing Laboratories (Beijing) Co., Ltd., under GLP principles consistent with FDA 21 CFR Part 58, the NMPA Good Laboratory Practice for Non-Clinical Laboratory Studies (Order No. 34, 2017), and the Center for Food and Drug Inspection of NMPA (CFDI) *Guideline for the Management of Bioanalytical Laboratories for Drug Clinical Trials* (September 2025), following approved study protocols and institutional standard operating procedures (study numbers IN-25-404-1MV, IFN-γ; IN-25-432-1MV, IL-2; IN-25-431-1MV, IL-4). Quality assurance unit oversight included facility audits and review of each final report against original data. Method-file versions were IN-25-404-1MV-V1/V2, IN-25-432-1MV-V1, and IN-25-431-1MV-V1.

### Ethics and human samples

Peripheral blood was collected from healthy adult donors into BD Vacutainer CPT tubes after informed consent under applicable ethics approvals. Whole blood was transported at controlled ambient temperature (10–30°C) with continuous temperature monitoring; a minimum interval of 3.5 h between collection and centrifugation was specified by protocol, and samples were held at room temperature and processed within 2 h of arrival. Both freshly isolated and cryopreserved PBMC were used as specified for each validation parameter. The study was approved by the Drug Clinical Trial Ethics Committee of Tianjin Occupational Disease Prevention and Treatment Hospital (approval No. 2024-044, dated 13 November 2024 and 3 November 2025) and the Clinical Trial Ethics Committee of Shijiazhuang Hospital of Traditional Chinese Medicine (approval No. 25/04/04-01, dated 15 April 2025).

### Key reagents and instruments

Human cytokine ELISpot was performed using CTL pre-coated strip plates and matched ImmunoSpot reagents: Human IFN-γ Single-Color Enzymatic ELISPOT Kit (catalog hIFNgp-2M/10 or hIFNgp-2M/2; lots I0312OHC and I0224OZK), ImmunoSpot® System T CELL SINGLE-COLOR ENZYMATIC Hu IL-2 (catalog hIL2p-2M/10; lot 21001OEW), and ImmunoSpot® System T CELL SINGLE-COLOR ENZYMATIC Hu IL-4 (catalog hIL4p-2M/10; lot 40917OHC), all from Cellular Technology Limited (CTL). Culture and wash reagents comprised CTL-Test™ Medium, CTL-Wash™, L-glutamine, and penicillin–streptomycin, prepared per manufacturer and SOP instructions.

#### Antigens

- **CEFX Ultra SuperStim Pool** (JPT Peptide Technologies; catalog PM-CEFX-2): used for IFN-γ and IL-2 assays. Lots 54003Gre01, 59276Gre01, and 60974Gre01 were used in the IFN-γ study and lot 62925Gre01 in the IL-2 study. Lyophilized peptide was reconstituted in DMSO and PBS (pH 7.4) to a 10 μM stock and diluted to a working concentration of 0.10 μM in CTL-TEST; aliquots were stored at −60 to −90°C.
- **CPI Antigen Pool** (Cellular Technology Limited; catalog **CTL-CPI-001**; lot 0703OEK, expiry 2027-07-03): used for IL-4 assays throughout all validation arms.

#### Controls

- Negative control: medium/diluent only.
- Positive control: GMP monoclonal anti-human CD3 antibody (OKT3; catalog GMP-MC0323, Acro Biosystems).
- Blank wells: diluent without PBMC.

PBMC counting used the CTL ImmunoSpot analyzer fluorescence cell-counting function within its validated dilution range (0.5×10□ to 5×10□ cells/mL). Spots were enumerated as spot-forming units (SFU) per well using the CTL ELISpot reader under locked counting settings defined in the study SOP.

### PBMC isolation, viability, plating, and cryopreservation

PBMC were isolated from CPT tubes by centrifugation (1800 ×g, 23°C, 30 min, minimum acceleration/deceleration), washed twice with CTL-Wash, and resuspended in CTL-TEST. Viability acceptance for analysis was ≥80%. Unless otherwise specified for linearity assessments, the standard plating density was 2×10□ PBMC/well. Cryopreservation used a controlled-rate freezing container, and thawing followed laboratory SOPs with anti-aggregate wash reagents where applicable.

### ELISpot procedure

For each cytokine assay, plates were prepared with Blank, Neg, CD3, and antigen (CEFX or CPI) wells (100 μL stimulant/control per well; Blank 200 μL diluent). After pre-warming of stimulants, PBMC suspensions were added and plates incubated at 37°C, 5% CO□ for the validated stimulation period:

| **Assay** | **Antigen** | **Stimulation time** |
|---|---|---|
| IFN-γ | CEFX | **24 h and 48 h** (both validated) |
| IL-2 | CEFX | **24 h** |
| IL-4 | CPI (CTL-CPI-001) | **48 h** |

After stimulation, wells were processed with detection antibody, streptavidin–alkaline phosphatase, and substrate according to the kit/SOP workflow. Dried plates were scanned and SFU/well recorded. Triplicate wells were used unless cell numbers constrained replication; mean SFU and %CV were calculated per SOP rules, including predefined outlier-well handling when triplicate %CV exceeded 40% (the two wells with the lowest %CV were retained).

### Validation parameters and acceptance criteria

Validation parameters and acceptance criteria were predefined in the study protocols and aligned with fit-for-purpose ELISpot practice and GCC recommendations. **Table 1** summarizes the design and acceptance criteria applied across the three assays.

**Table 1.**
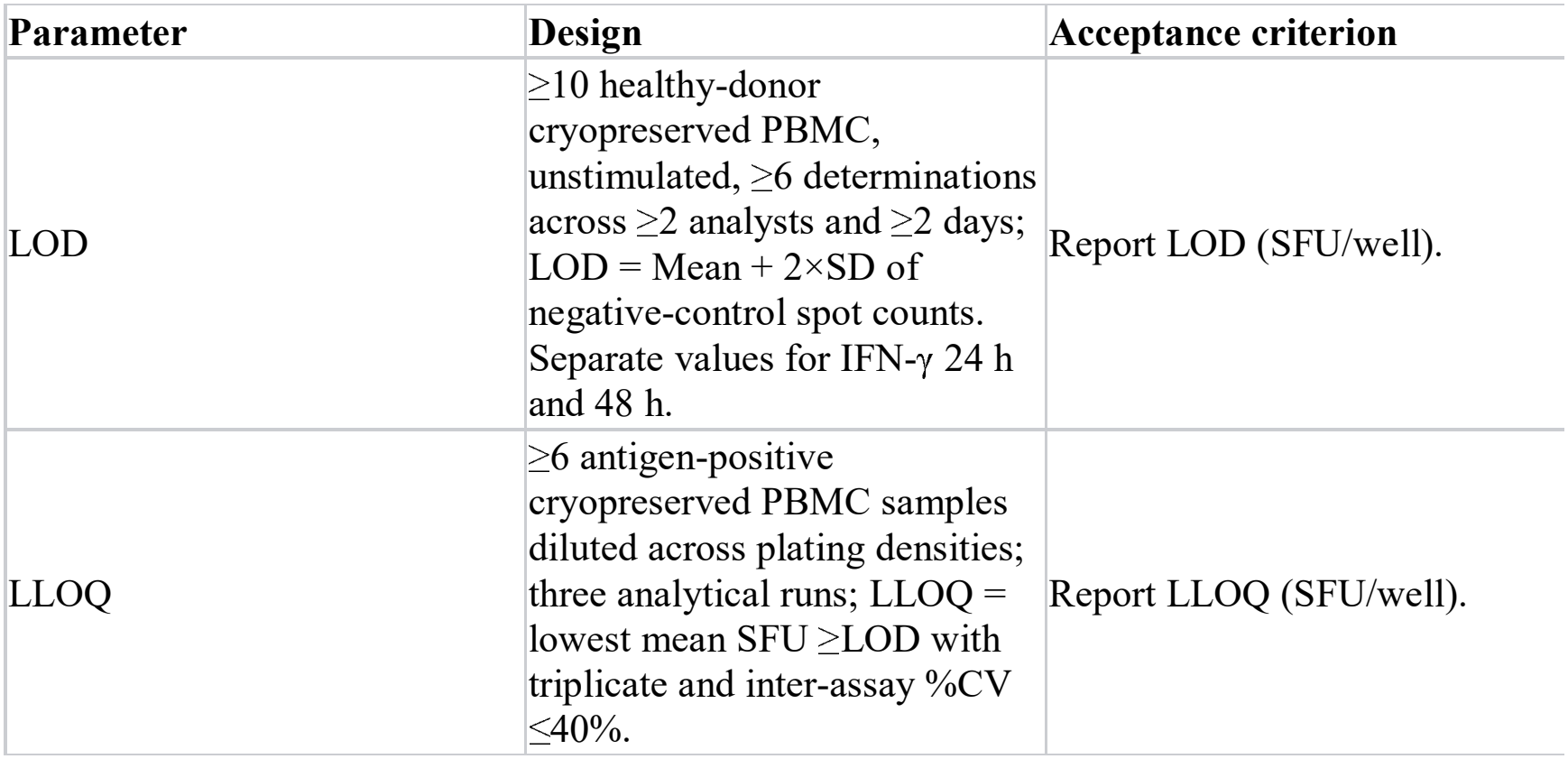
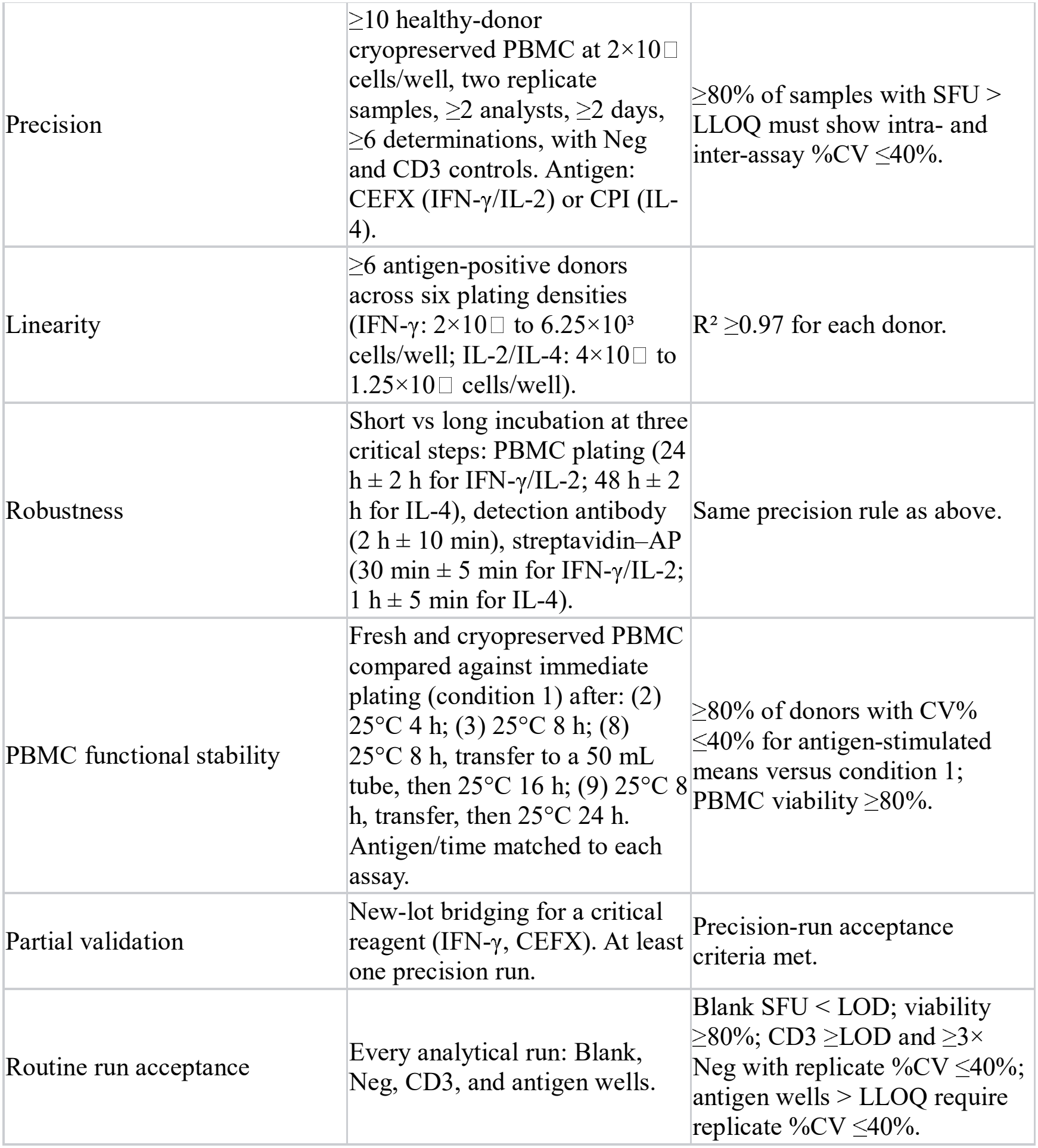
Validation design and predefined acceptance criteria.

### Data analysis

Descriptive statistics (mean, SD, %CV) and linear regression (R^2^) were calculated as defined in the protocols. LOD used Mean + 2×SD. For stability comparisons, %CV was computed between the antigen-stimulated mean under each delayed condition and the mean under condition 1. Results are reported as SFU/well unless otherwise stated. Statistical analysis was performed with Microsoft Excel.

## Results

### Overall validation outcome

All three ELISpot methods met the predefined acceptance criteria for LOD, LLOQ, precision, linearity, robustness, and fresh-sample PBMC functional stability, and were concluded fit for the intended use.

Key numerical outcomes are summarized in **Table 2**.

**Table 2.**
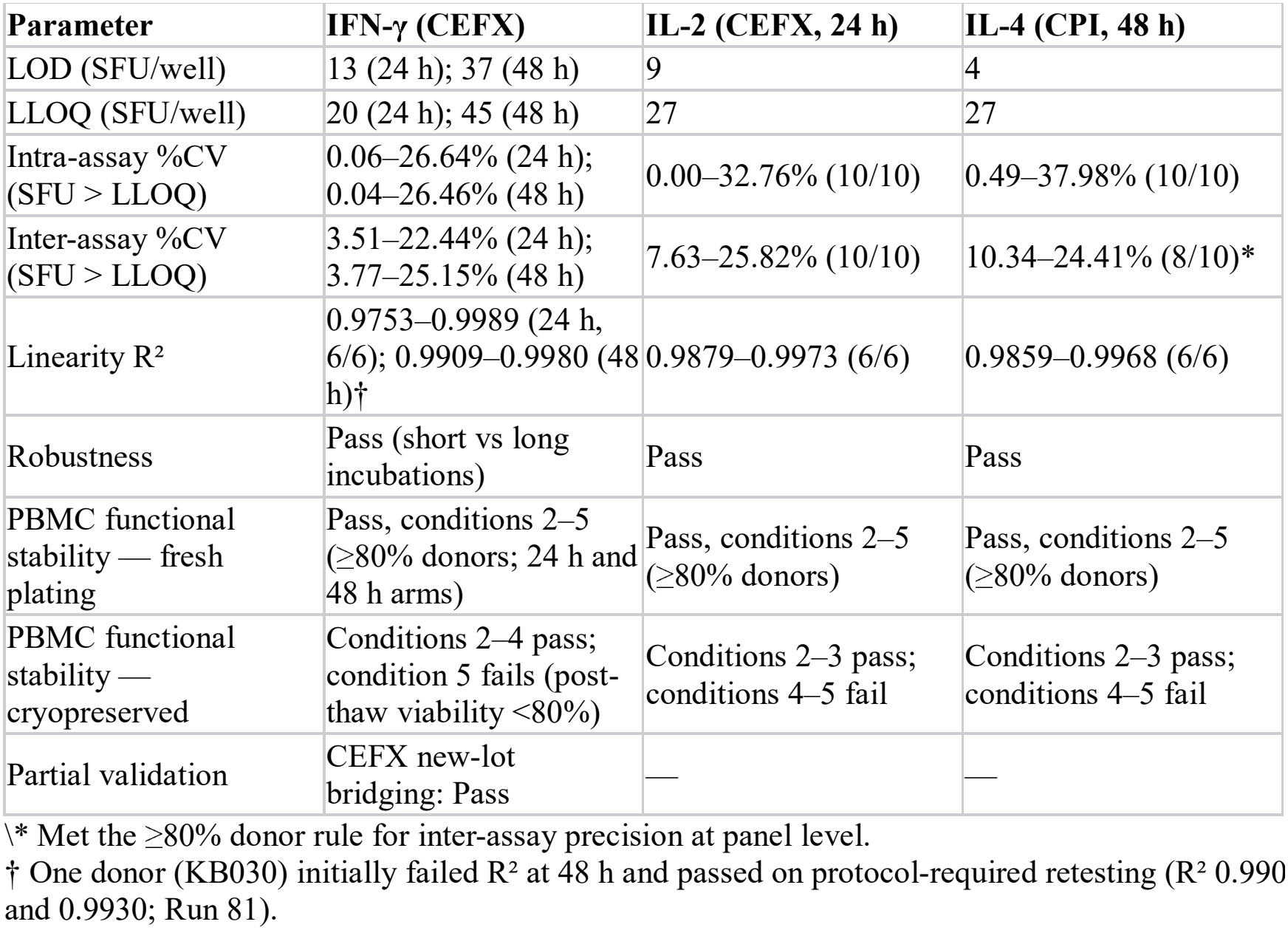
Summary of validated performance (values from GLP validation conclusions)

| **Parameter** | **IFN-γ (CEFX)** | **IL-2 (CEFX, 24 h)** | **IL-4 (CPI, 48 h)** |
|---|---|---|---|
| LOD (SFU/well) | 13 (24 h); 37 (48 h) | 9 | 4 |
| LLOQ (SFU/well) | 20 (24 h); 45 (48 h) | 27 | 27 |
| Intra-assay %CV (SFU > LLOQ) | 0.06–26.64% (24 h);<br>0.04–26.46% (48 h) | 0.00–32.76% (10/10) | 0.49–37.98% (10/10) |
| Inter-assay %CV (SFU > LLOQ) | 3.51–22.44% (24 h);<br>3.77–25.15% (48 h) | 7.63–25.82% (10/10) | 10.34–24.41% (8/10)* |
| Linearity R² | 0.9753–0.9989 (24 h, 6/6); 0.9909–0.9980 (48 h)† | 0.9879–0.9973 (6/6) | 0.9859–0.9968 (6/6) |
| Robustness | Pass (short vs long incubations) | Pass | Pass |
| PBMC functional stability — fresh plating | Pass, conditions 2–5 (≥80% donors; 24 h and 48 h arms) | Pass, conditions 2–5 (≥80% donors) | Pass, conditions 2–5 (≥80% donors) |
| PBMC functional stability — cryopreserved | Conditions 2–4 pass; condition 5 fails (post-thaw viability <80%) | Conditions 2–3 pass; conditions 4–5 fail | Conditions 2–3 pass; conditions 4–5 fail |
| Partial validation | CEFX new-lot bridging: Pass | — | — |

\\\* Met the ≥80% donor rule for inter-assay precision at panel level.
† One donor (KB030) initially failed R² at 48 h and passed on protocol-required retesting (R² 0.990[illegible] and 0.9930; Run 81).

### LOD and LLOQ

The IFN-γ LOD/LLOQ differed by stimulation duration. Both thresholds were higher at 48 h (LOD 37; LLOQ 45 SFU/well) than at 24 h (LOD 13; LLOQ 20 SFU/well), a pattern consistent with increased background and variance during longer culture. IL-2 LOD/LLOQ were 9 and 27 SFU/well. Notably, IL-4 showed the lowest LOD (4 SFU/well), with LLOQ 27 SFU/well under CPI 48 h stimulation. Blank wells met the Blank < LOD criterion across all validation batches (85, 40, and 53 analytical runs for IFN-γ, IL-2, and IL-4, respectively; Supplementary Tables).

### Precision

For CEFX-stimulated IFN-γ wells above LLOQ, intra-assay %CV ranges were 0.06–26.64% (24 h) and 0.04–26.46% (48 h). Inter-assay %CV ranges were 3.51–22.44% (24 h) and 3.77–25.15% (48 h). Both met the ≥80% / ≤40% acceptance rule. IL-2 CEFX precision passed for 10/10 donors (intra 0.00–32.76%; inter 7.63–25.82%). IL-4 CPI precision passed at the panel level under the same rule (intra 0.49–37.98% for 10/10 donors; inter 10.34–24.41% for 8/10 donors). Robustness arms likewise maintained precision within acceptance after short versus long critical incubations: IFN-γ intra 0.00– 19.85% (24 h) and 0.27–34.14% (48 h), inter 2.08–32.47% and 3.81–19.86%; IL-2 intra 2.32–17.10% and inter 5.22–24.41% (10/10); IL-4 intra 0.61–24.73% and inter 2.64–22.46% (10/10).

### Linearity

SFU increased linearly with plated PBMC density for all cytokines. IFN-γ R^2^ values were 0.9753– 0.9989 at 24 h (6/6 donors) and, after protocol-required retesting of one 48 h donor, 0.9909–0.9980 at 48 h. IL-2 R^2^ was 0.9879–0.9973 (6/6) and IL-4 R^2^ was 0.9859–0.9968 (6/6). All met the R^2^ ≥0.97 criterion across the six-density range.

### Robustness

Varying PBMC plating, detection antibody, and streptavidin–AP incubation times within predefined short/long windows did not compromise assay performance; precision acceptance was retained for antigen responses above LLOQ for IFN-γ (CEFX), IL-2 (CEFX), and IL-4 (CPI). One protocol deviation in the IFN-γ study (D01; streptavidin–AP incubation shorter than the specified short window in Runs 9–10) was assessed in dedicated runs: intra-assay %CV 0.60–25.71% and inter-assay %CV 10.30–19.63%, within acceptance, and the deviation was judged not to affect the reported results (Supplementary Table S11).

### Pre-analytical PBMC functional stability

Fresh-plating arms passed for all three assays (Figure 4). Relative to immediate plating (condition 1), antigen-stimulated means under conditions 2–5 met the CV% ≤40% criterion in ≥80% of donors. For IFN-γ 24 h CEFX the counts were 11/12, 12/12, 12/12, and 11/12 donors; IFN-γ 48 h CEFX 8/10, 9/10, 9/10, and 9/10; IL-2 24 h CEFX 12/12, 12/12, 12/12, and 11/12; and IL-4 48 h CPI 12/12, 12/12, 12/12, and 11/12, respectively. Thus, freshly plated responses remained within acceptance even after 8 h of ambient holding followed by transfer and a further 24 h hold before plating. Cryopreserved arms showed a narrower, assay-dependent window. For IFN-γ, conditions 2, 3, and 4 passed (9/11, 11/12, 9/11 donors at 24 h and 8/10, 9/10, 8/10 at 48 h), whereas condition 5 failed: seven donors showed post-thaw viability <80% and the arm was terminated, with the same outcome on repeat testing (Runs 77–78 and 80). For IL-2, conditions 2 and 3 passed (12/12 donors each), condition 4 failed (only 5/12 donors within CV% ≤40%; 7/12 exceeded 40%), and condition 5 failed because post-thaw viability was <80% in 11 donors; the single donor with acceptable viability failed the CD3 run-acceptance criterion (Run 40). For IL-4, conditions 2 and 3 passed (12/12 and 10/12 donors), condition 4 failed (5/12 donors within criterion; 7/12 exceeded 40%), and condition 5 failed because all 12 donors had post-thaw viability <80% and testing was terminated (Runs 54–56). A complete pass/fail matrix for fresh and cryopreserved arms is provided in Supplementary Table S38. Taken together, the data support ambient holding of whole blood/PBMC for up to 8 h plus a further 24 h when samples are plated fresh. They further indicate that cryopreservation should be performed within 8 h of the initial hold, and that prolonged delayed processing before freezing is not supported by this validation package.

**Figure 1.**
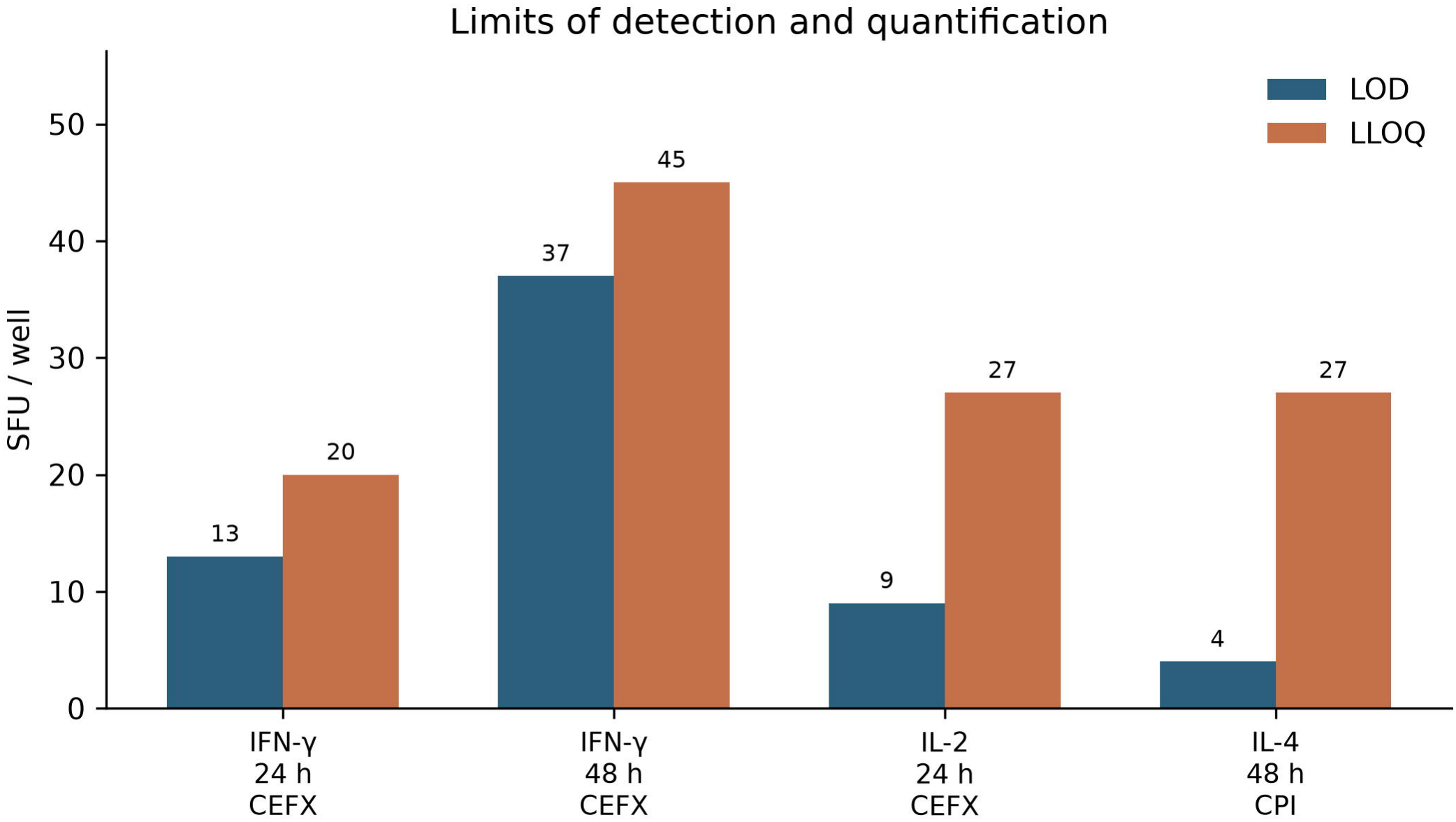
Limits of detection and quantification. LOD and LLOQ (SFU/well) for IFN-γ ELISpot (CEFX, 24 h and 48 h), IL-2 ELISpot (CEFX, 24 h), and IL-4 ELISpot (CPI Antigen Pool CTL-CPI-001, 48 h). Values are taken from the GLP validation conclusions (IN-25-404/432/431-1MV).

**Figure 2.**
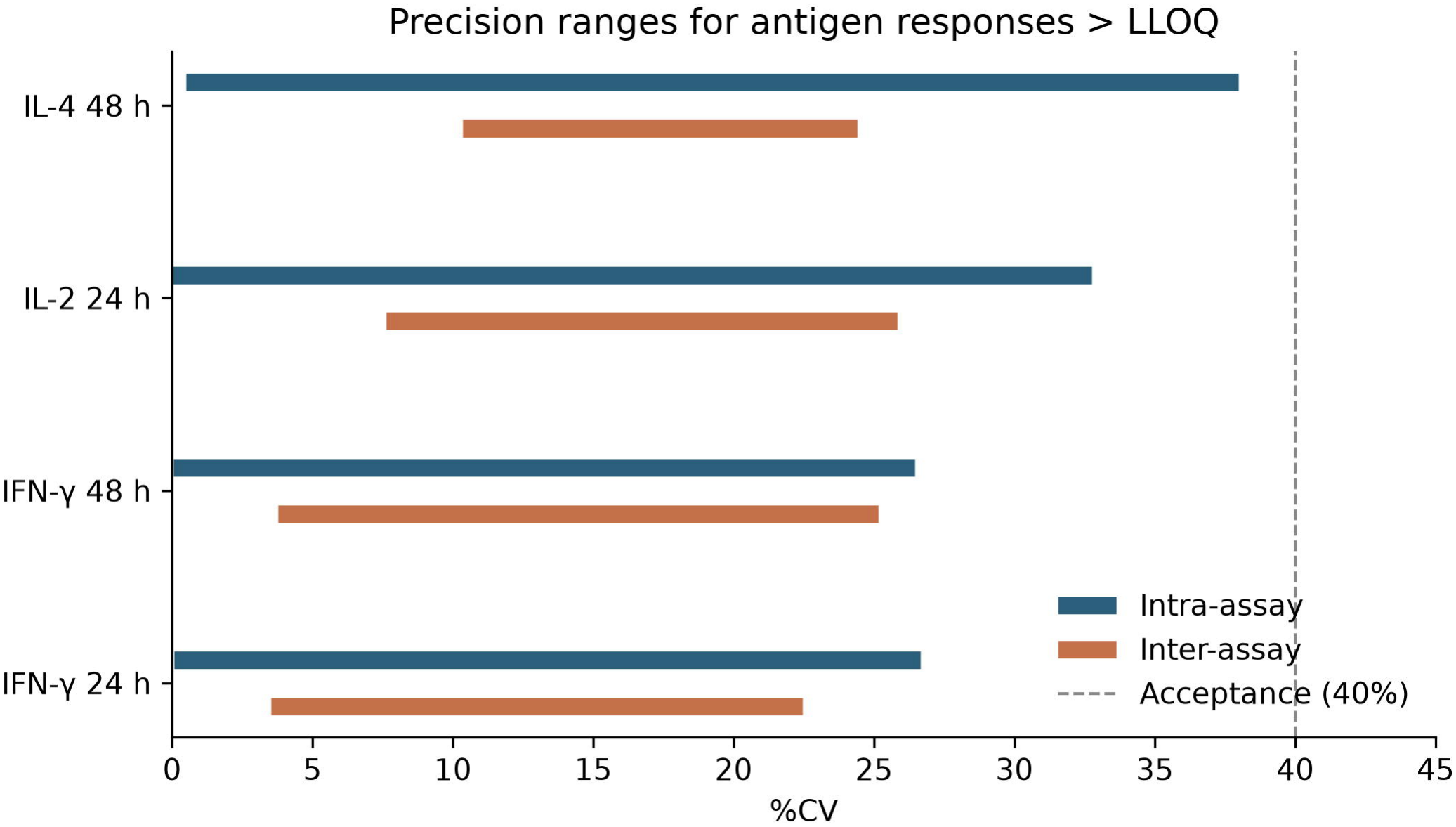
Precision ranges for antigen responses above LLOQ. Horizontal bars show observed intra-assay and inter-assay %CV ranges for CEFX- or CPI-stimulated wells with SFU > LLOQ. The dashed line marks the 40% acceptance threshold used with the ≥80% donor/sample rule.

**Figure 3.**
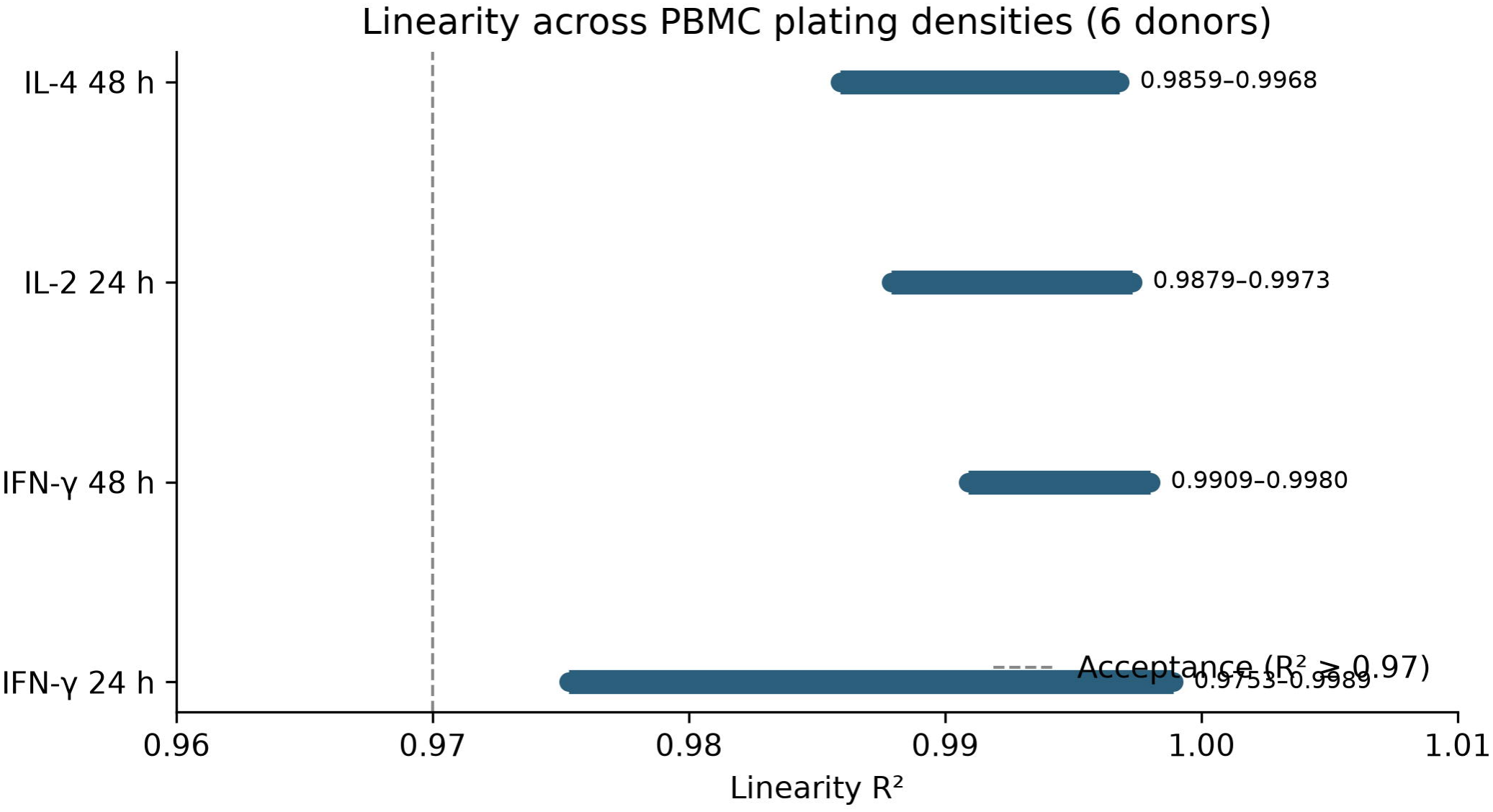
Linearity of SFU versus PBMC plating density. Min–max R^2^ across six antigen-positive donors per assay. Acceptance criterion: R^2^ ≥0.97 for each donor. The IFN-γ 48 h result includes the protocol-required retest of donor KB030. Representative individual regression plots are provided in Supplementary Figures.

**Figure 4.**
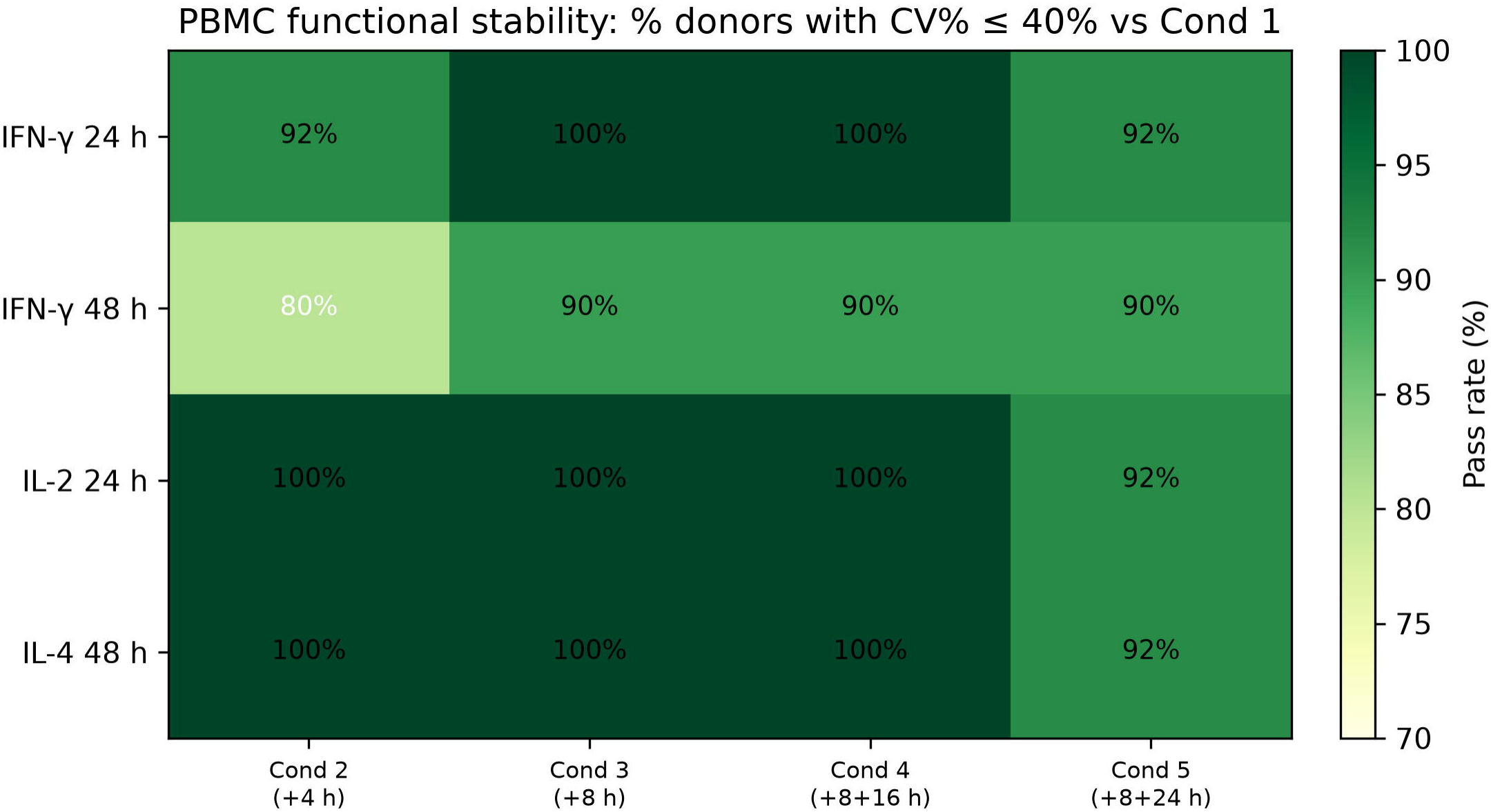
Pre-analytical PBMC functional stability — fresh-plating arm. Percentage of donors meeting CV% ≤40% for antigen-stimulated means under delayed-processing conditions 2–5 versus immediate plating (condition 1). Conditions: (2) 25°C 4 h; (3) 25°C 8 h; (4) 8 h then transfer to a 50 mL tube + 16 h; (5) 8 h then transfer + 24 h. Acceptance required ≥80% of donors. **This figure displays the fresh-plating arm only**; cryopreserved-arm outcomes, including the failed conditions, are reported in the Results section and Supplementary Table S38.

### Partial validation of critical reagents

Dedicated IFN-γ runs supported bridging to a new CEFX Ultra SuperStim Pool lot (60974Gre01; Runs 82–85), meeting precision-run acceptance criteria (Supplementary Table S17). These data support critical-reagent continuity for ongoing clinical testing.

## Discussion

ELISpot remains a cornerstone method for quantifying antigen-specific cellular immunity in clinical development. Although IFN-γ-centric assays dominate vaccine and cell-therapy immunomonitoring, parallel assessment of IL-2 and IL-4 expands biological coverage across Th1-associated activation and Th2-axis responses. Recent formal validations of dual-color IFN-γ/IL-4 ELISpot and IFN-γ/IL-2 FluoroSpot assays demonstrate that multi-cytokine spot formats can meet bioanalytical acceptance criteria for clinical use (8,9), and continued protocol optimization is refining sensitivity for low-frequency responses (10,16). The present work complements those reports by delivering a GLP-aligned, three-assay single-color panel validated under one laboratory quality system, with explicit operational emphasis on pre-analytical PBMC stability and critical-reagent management. We intentionally paired CEFX Ultra SuperStim Pool with IFN-γ and IL-2, and CPI Antigen Pool (CTL-CPI-001) with IL-4. CEFX provides a broad recall peptide stimulus that elicits robust type 1 cytokine responses in healthy-donor PBMC, enabling sensitive precision and linearity assessments. CPI was selected for IL-4 because it reproducibly supported Th2-axis spot formation under the 48 h stimulation regimen used throughout IL-4 validation. Using assay-matched recall antigens is consistent with fit-for-purpose ELISpot practice: the validation antigen need not be the eventual clinical study antigen, but must generate measurable, physiologically relevant responses for method characterization (11-13). When the panel is later applied to product-specific peptides, partial validation or bridging can be executed against the acceptance framework established here. Stimulation duration materially affected IFN-γ performance characteristics. Both 24 h and 48 h CEFX regimens met precision and linearity criteria, but LOD/LLOQ were higher at 48 h (37/45 SFU/well) than at 24 h (13/20 SFU/well). This pattern is consistent with increased background and/or variance during longer culture and should guide protocol selection: 24 h may be preferable when maximizing low-level sensitivity is critical, whereas 48 h may be retained when the stimulus is not a defined peptide pool but long peptides or proteins, or in other situations favoring longer stimulation. IL-2 was validated at 24 h and IL-4 at 48 h, reflecting cytokine-specific response kinetics established during method development.

Importantly, precision acceptance (≥80% of samples with intra- and inter-assay %CV ≤40% above LLOQ) aligns with consensus recommendations for cell-based ELISpot assays, which typically tolerate higher imprecision than ligand-binding assays (11,12). All three methods satisfied this framework. Surprisingly, IL-4 inter-assay precision passed at the panel level even though only 8/10 donors met the ≤40% criterion, which illustrates why population-level (≥80%) rules are preferred over requiring every individual donor to pass. Linearity R^2^ ≥0.97 across six plating densities supports quantitative interpretation of SFU as proportional to responder-cell input within the validated range. A distinctive contribution of this validation package is the pre-analytical stability design. Its most decision-relevant finding, however, is a boundary rather than a blanket pass. Freshly plated PBMC retained antigen-specific responses within acceptance across all five handling conditions for all three cytokines, supporting ambient shipment windows as long as 8 h followed by a further 24 h before plating. Cryopreserved material was less forgiving. IFN-γ tolerated cryopreservation after up to 8+16 h of delayed processing but failed at 8+24 h, whereas IL-2 and IL-4 failed at both 8+16 h and 8+24 h. The dominant failure mode was post-thaw viability below the 80% acceptance threshold, with a secondary mode of excessive response variability after prolonged ambient holding before freezing. These observations are consistent with the recognized sensitivity of cryopreserved PBMC to processing delay. They also agree with reports that viability loss and functional decline, rather than analytical imprecision, limit delayed-processing workflows (3,5,10). For multicenter trials the practical implication is explicit. If samples are to be cryopreserved for batched analysis, freezing should occur within 8 h of the initial ambient hold. If longer delays are unavoidable, fresh plating within the validated window is the supported option. Reporting this boundary rather than collapsing all arms into a single “pass” strengthens, rather than weakens, the claim of fit-for-purpose validation, because it defines the conditions under which the assay is actually qualified.

Combined with robustness testing of critical incubation windows and CEFX new-lot bridging for IFN-γ, these data address operational failure modes that purely analytical validations often omit. GCC recommendations explicitly highlight these modes as required SOP content (11). The three assays are single-color rather than multiplex, which increases PBMC demand when all three cytokines are required from one sample. FluoroSpot or dual-color formats may be preferable when cell numbers are limiting (8,9). This limitation is partially offset by the validated linearity ranges, which permit reduced plating densities when cell yield is constrained.

Several additional limitations should be noted. Validation used healthy-donor PBMC and recall antigen pools rather than disease- or product-specific cohorts; clinical cut-point strategies and antigen-specific positivity algorithms must be defined in study-specific contexts. Relative accuracy against an independent orthogonal method was not a primary objective. We instead emphasized precision, sensitivity limits, linearity, robustness, and stability for fit-for-purpose deployment. Inter-laboratory concordance was not assessed in this package, unlike the inter-laboratory comparison reported for dual-color FluoroSpot (9), and would strengthen future harmonization work. Finally, the cryopreserved 8+24 h failure was driven by viability rather than by cytokine response per se, so alternative cryopreservation media or thawing procedures might extend that window; such changes would require dedicated validation.

In summary, the validated IFN-γ/IL-2 (CEFX) and IL-4 (CPI) ELISpot methods provide a practical Th1/Th2 cellular immunity platform suitable for regulated clinical immunomonitoring when applied within the reported acceptance criteria and the explicitly defined sample-handling boundaries.

## Conclusion

We completed GLP-aligned, fit-for-purpose bioanalytical validation of three human PBMC ELISpot assays: IFN-γ and IL-2 with CEFX Ultra SuperStim Pool (24 h, with an additional validated 48 h IFN-γ regimen) and IL-4 with CPI Antigen Pool (CTL-CPI-001, 48 h). LOD, LLOQ, precision, linearity, robustness, and fresh-sample PBMC functional stability met predefined acceptance criteria consistent with Global CRO Council recommendations, and critical-reagent new-lot bridging was qualified for the IFN-γ assay. Cryopreserved-sample stability was qualified only within shorter delayed-processing windows, defining an operational boundary for clinical sample logistics. Within these acceptance criteria and handling boundaries, the three-cytokine panel is fit-for-purpose for quantitative cellular immunity monitoring in regulated clinical trials.

## Data Availability

De-identified summary validation datasets supporting the conclusions are provided in the Supplementary Material (see the index for the supplementary tables and machine-readable CSV summaries). Original GLP raw data, protocols, and reports are archived at the testing facility according to institutional SOPs and applicable retention requirements. Access requests may be directed to the corresponding author, subject to ethical and contractual constraints.

## Data Availability Statement

De-identified summary validation datasets supporting the conclusions of this study are provided in the Supplementary Material. Original GLP raw data, protocols and reports are archived at JOINN Medical Testing Laboratories (Beijing) Co., Ltd. in accordance with institutional SOPs and applicable retention requirements. Access requests may be directed to the corresponding author (Kaitong Li), subject to ethical and contractual constraints.

## Ethics Statement

Studies involving human blood donors were conducted in accordance with the Declaration of Helsinki and applicable local regulations. Donors provided informed consent. Ethics approval was granted by the Drug Clinical Trial Ethics Committee of Tianjin Occupational Disease Prevention and Treatment Hospital (approval No. 2024-044, dated 13 November 2024 and 3 November 2025) and the Clinical Trial Ethics Committee of Shijiazhuang Hospital of Traditional Chinese Medicine (approval No. 25/04/04-01, dated 15 April 2025).

## Author Contributions

M. Tang and P. Guo supervised the experimental work and established the assay methodology. X. Zhai, H. Cao, N. Zhang and P. Fan performed the validation experiments and acquired the data. L. Jiang was responsible for project administration and coordination. Z. Cui handled ethics approval, informed-consent documentation and regulatory compliance. H. Wang and Z. Wang performed independent quality assurance (QA) oversight and GLP compliance auditing. K. Li conceived and supervised the study, drafted the manuscript and carried out the final scientific review. All authors read and approved the final manuscript and agree to be accountable for all aspects of the work.

## Acknowledgments

We thank the donors and laboratory staff who contributed to sample collection and assay execution. Generative AI tools were used only for the language polishing of the manuscript text, and all the analyses, the scientific content and the conclusions were produced and verified by the authors.

