## Supplementary material for "Fit-for-purpose bioanalytical validation of a three-cytokine human PBMC ELISpot panel (IFN-γ, IL-2, IL-4) using CEFX and CPI antigen pools for clinical trial immunomonitoring under a GLP-aligned quality system": Supplementary_Material_AAPS.docx

### 1. File list

| **Supp ID** | **Content** | **Source** | **File** |
| --- | --- | --- | --- |
| S01–S02 | LOD raw data, 24 h / 48 h CEFX | IFN-γ annexes 1–2 | S_tables/Table_S01.csv … S02.csv |
| S03–S04 | LLOQ raw data, 24 h / 48 h | IFN-γ annexes 3–4 | Table_S03.csv … S04.csv |
| S05–S06 | Blank QC / PBMC run QC | IFN-γ annexes 5–6 | Table_S05.csv … S06.csv |
| S07–S08 | Precision, 24 h / 48 h | IFN-γ annexes 7–8 | Table_S07.csv … S08.csv |
| S09–S11 | Robustness incl. deviation D01 | IFN-γ annexes 9–11 | Table_S09.csv … S11.csv |
| S12–S13 | PBMC functional stability, 24 h / 48 h | IFN-γ annexes 12–13 | Table_S12.csv … S13.csv |
| S14–S16 | Linearity, 24 h / 48 h / KB030 retest | IFN-γ annexes 14–16 | Table_S14.csv … S16.csv |
| S17 | Partial validation (CEFX new-lot bridging) | IFN-γ annex 17 | Table_S17.csv |
| S18 | LOD | IL-2 annex 1 | Table_S18.csv |
| S19 | LLOQ | IL-2 annex 2 | Table_S19.csv |
| S20–S21 | Blank QC / PBMC QC | IL-2 annexes 3–4 | Table_S20.csv … S21.csv |
| S22 | Precision | IL-2 annex 5 | Table_S22.csv |
| S23 | Robustness | IL-2 annex 6 | Table_S23.csv |
| S24 | PBMC functional stability (fresh + cryopreserved) | IL-2 annex 7 | Table_S24.csv |
| S25 | Linearity | IL-2 annex 8 | Table_S25.csv |
| S27 | LOD | IL-4 annex 1 | Table_S27.csv |
| S28 | LLOQ | IL-4 annex 2 | Table_S28.csv |
| S29–S30 | Blank QC / PBMC QC | IL-4 annexes 3–4 | Table_S29.csv … S30.csv |
| S31 | Precision | IL-4 annex 5 | Table_S31.csv |
| S32 | Robustness | IL-4 annex 6 | Table_S32.csv |
| S33 | PBMC functional stability (fresh + cryopreserved) | IL-4 annex 7 | Table_S33.csv |
| S34 | Linearity | IL-4 annex 8 | Table_S34.csv |
| S35 | Supplementary Methods (condensed SOP flow) | Report methods sections | this document, §2 |
| S36 | Unified validation acceptance criteria | Report §2.7 / main-text Table 1 | this document, §3 |
| S37 | Critical reagents, catalog numbers and lots | Report key-reagent tables | this document, §4 |
| S38 | PBMC stability pass/fail matrix (fresh + cryopreserved) | Report stability conclusions | Table_S38_stability_matrix.csv |

Supplementary IDs, source annexes and data-row counts are mapped in _INDEX.csv. All CSVs are UTF-8 with BOM and entirely in English; tabular data were exported directly from the source reports without recomputation. Figure 2 and Figure 4 data are provided as Figure2_precision_ranges.csv and Figure4_stability_pass.csv.

### 2. S35 — Supplementary Methods (brief summary)

Detailed procedures are defined in the site standard operating procedures (SOPs) and the study protocol. In brief, whole blood was collected in BD Vacutainer CPT tubes, transported at 10–30 °C with continuous temperature monitoring, and processed to PBMC by density-gradient centrifugation. PBMC were counted, assessed for viability, and plated at a defined cell density. Cells were stimulated with recall antigen peptide pools (CEFX for IFN-γ and IL-2; CPI for IL-4) for 24 or 48 h, and ELISpot plates were developed and read according to the validated assay procedure. Critical reagents, acceptance criteria and stability conclusions are summarised in S36–S38 below.

### 3. S36 — Unified validation acceptance criteria

| **Parameter** | **Design** | **Acceptance** |
| --- | --- | --- |
| LOD | ≥10 donors, ≥6 determinations, ≥2 analysts, ≥2 days | LOD = Mean + 2×SD; report value |
| LLOQ | ≥6 antigen-positive donors, 3 runs, serial plating densities | Lowest mean SFU ≥LOD with triplicate and inter-assay %CV ≤40% |
| Precision | ≥10 donors, 2 replicates, ≥2 analysts, ≥2 days, ≥6 determinations | ≥80% of samples above LLOQ with intra- and inter-assay %CV ≤40% |
| Linearity | ≥6 donors, 6 plating densities | R² ≥0.97 per donor |
| Robustness | Short vs long incubation at PBMC plating, detection antibody, streptavidin–AP steps | Same precision rule |
| PBMC functional stability | Conditions 1–5; fresh and cryopreserved arms; antigen/time matched to assay | ≥80% donors with CV% ≤40% vs condition 1; viability ≥80% |
| Partial validation | ≥1 precision run per reagent change | Precision-run criteria met |
| Routine run | Blank, Neg, CD3, antigen per plate | Blank < LOD; viability ≥80%; CD3 ≥LOD and ≥3× Neg; antigen >LLOQ requires %CV ≤40% |

### 4. S37 — Critical reagents, catalog numbers and lots

| **Reagent** | **Catalog** | **Lot(s)** | **Manufacturer** |
| --- | --- | --- | --- |
| Human IFN-γ Single-Color Enzymatic ELISPOT Kit | hIFNgp-2M/10; hIFNgp-2M/2 | I0312OHC; I0224OZK | CTL |
| ImmunoSpot® System T CELL SINGLE-COLOR ENZYMATIC Hu IL-2 | hIL2p-2M/10 | 21001OEW | CTL |
| ImmunoSpot® System T CELL SINGLE-COLOR ENZYMATIC Hu IL-4 | hIL4p-2M/10 | 40917OHC | CTL |
| CEFX Ultra SuperStim Pool | PM-CEFX-2 | 54003Gre01, 59276Gre01, 60974Gre01 (IFN-γ); 62925Gre01 (IL-2) | JPT |
| CPI Antigen Pool | CTL-CPI-001 | 0703OEK (expiry 2027-07-03) | CTL |
| GMP monoclonal anti-human CD3 (OKT3) | GMP-MC0323 | C15dC2-G002-228JF1-228K | Acro Biosystems |
| CTL-Test™ Medium / CTL-Wash™ | kit supplied | — | CTL |

### 5. S38 — PBMC functional stability pass/fail matrix

See Table_S38_stability_matrix.csv for the complete machine-readable matrix (32 rows: 4 assay arms × 2 sample arms × 4 conditions). Summary:

| **Assay** | **Fresh-plating arm** | **Cryopreserved arm** |
| --- | --- | --- |
| IFN-γ 24 h CEFX | Pass, conditions 2–5 (11/12, 12/12, 12/12, 11/12) | Conditions 2–4 pass (9/11, 11/12, 9/11); condition 5 fails (post-thaw viability <80%) |
| IFN-γ 48 h CEFX | Pass, conditions 2–5 (8/10, 9/10, 9/10, 9/10) | Conditions 2–4 pass (8/10, 9/10, 8/10); condition 5 fails (post-thaw viability <80%) |
| IL-2 24 h CEFX | Pass, conditions 2–5 (12/12, 12/12, 12/12, 11/12) | Conditions 2–3 pass (12/12, 12/12); condition 4 fails (5/12); condition 5 fails (viability) |
| IL-4 48 h CPI | Pass, conditions 2–5 (12/12, 12/12, 12/12, 11/12) | Conditions 2–3 pass (12/12, 10/12); condition 4 fails (5/12); condition 5 fails (12/12 viability <80%) |

Condition definitions: (1) immediate plating; (2) 25°C 4 h; (3) 25°C 8 h; (4) 25°C 8 h, transfer to a 50 mL tube, then 25°C 16 h; (5) 25°C 8 h, transfer, then 25°C 24 h. Acceptance required ≥80% of donors with CV% ≤40% versus condition 1.
